# Selenium supplementation and clinical outcomes: an umbrella review protocol

**DOI:** 10.1101/2023.05.26.23290575

**Authors:** Adaobi Uchenna Mosanya, Ifeoma Umeh, Abba Khalid Abdullahi, Blessing Onyinye Ukoha-Kalu, Maxwell Ogochukwu Adibe, Carmen Sanmartin Grijalba

## Abstract

**Background:** Selenium is a trace element essential for the normal functions of different human body systems and its deficiency has been associated with different disease states. In recent years, many systematic reviews of randomized clinical trials in humans have demonstrated various properties of selenium such as antioxidant, anti-inflammatory, increased immunity, blocking tumour invasion and metastasis in various pathological conditions. Therefore, there is need to summarize these recent findings in one single paper to facilitate the decision making of clinicians and policy makers regarding the inclusion of selenium supplementation in routine clinical practice, treatment guidelines and essential medicine list. This study would guide researchers towards future research and design of clinical trials.

**Objectives:** The objective of this umbrella review is to assess the clinical outcomes of selenium supplementation in different disease states and to determine its therapeutic implications.

**Eligibility criteria:** Only systematic reviews of randomized clinical trials reporting clinical outcomes after the use of selenium alone as a supplement in the management of diseases will be included. While primary studies, systematic reviews that involved the administration of selenium in combination with other trace elements for the prevention or treatment of diseases will be excluded.

**Methods:** The following databases will be searched by three independent reviewers: MEDLINE, PUBMED, EMBASE, COCHRANE database of systematic reviews, CINAHL, JBI Evidence synthesis, EPISTEMONIKOS, SCOPUS, Web of Science and TRIP PRO. Unpublished reviews will be searched using ProQuest for dissertations and Theses, Canadian Agency for drugs and Technologies in Health (CADTH) database of Grey matters and GOOGLE scholar. Systematic reviews published in the last 10 years (2013 to 2023). There will be no language restriction. The services of a translator will be employed if studies in other languages were found. Selection of reviews and data extraction will be done by 3 independent reviewers. Summary of findings will be presented in tables accompanied by texts where necessary.

**Strengths and limitations of this study:** This is the first umbrella review on the use of selenium supplements for disease management. Secondly, the findings from this study would facilitate the decision making of clinicians and policy makers regarding the inclusion of selenium supplementation in routine clinical practice, treatment guidelines and essential medicine list. Nevertheless, a major limitation of this umbrella review would be a reduced scope of eligible studies because only systematic reviews/ or meta-analysis that included randomized clinical trials will be considered.

## Introduction

Selenium is a trace element essential for the normal functions of different human body systems because it forms part of selenium-glutathione peroxidase enzymes as a co-factor^1^. Some of its roles are critical in protecting against harm caused by oxidative stress, reproductive functions, metabolism of thyroid hormones as well as immunity against infections^2,3^ Its deficiency has been associated with the risk of developing different diseases such as Polycystic ovaries (PCOS)^4,5^, Cancer^6^, Tuberculosis^7^ Chronic liver diseases such as hepatitis, cirrhosis and liver cancer^8^. In recent years, many systematic reviews of randomized clinical trials in humans have demonstrated various properties of selenium such as antioxidant, anti-inflammatory, increased immunity, blocking tumour invasion and metastasis in various pathological conditions^9,10^.

Intravenous Selenium supplementation was found to be beneficial to acute respiratory diseases by reducing the inflammatory cytokines among other mechanisms thereby shortening the number of days spent in the hospital and mortality rate^11^ Other studies have identified its usefulness in trauma patients where those who received selenium had lesser length of stay both in the hospital and in the intensive care unit (ICU)^12^ Similar results was found in patients with severe sepsis or shock by reducing their hospital length of stay^13^ It was found to improve the glutathione peroxidase levels and activity as well as some cognitive tests in patients with Alzheimer’s disease and cognitive impairment^14^

Therefore, there is need to summarize these recent findings in one single paper to facilitate the decision making of clinicians and policy makers regarding the inclusion of selenium supplementation in routine clinical practice, treatment guidelines and essential medicine list. This study would also guide researchers towards future research and design of clinical trials. The objectives of this umbrella review is to assess the clinical outcomes of selenium supplementation to measure its impact in different disease states and to determine its therapeutic implications.

An exploratory search of the following databases: PROSPERO, PUBMED, COCHRANE database of Systematic Reviews, and JBI Evidence Synthesis was done on February 28, 2023 and there was no published nor on-going umbrella review on the topic.

### Review questions

What are the specific clinical outcomes produced by the administration of selenium supplements?

Do these clinical outcomes give sufficient evidence for routine therapeutic use of selenium in patients?

### Objectives

The objective of this umbrella review is to assess the clinical outcomes of selenium supplementation in different disease states and to determine its therapeutic implications.

### Eligibility criteria

The guideline provided by JBI for umbrella review protocol was followed for the eligibility criteria.

#### Participants

This review will examine systematic reviews that included the use of selenium supplementation alone in patients suffering from diseases under investigation. Systematic reviews that included the use of selenium supplementation in healthy subjects will be excluded.

#### Phenomena of interest

Systematic reviews of controlled trials that involved Selenium supplementation alone will be included. The dose of Selenium supplementation for the patients suffering from the different disease states under investigation will be identified. While systematic reviews of controlled trials that did not involve the use of Selenium supplementation alone will be excluded. Systematic reviews that involved the administration of selenium in combination with other trace elements for the prevention or treatment of diseases will be excluded.

#### Outcomes

The clinical outcomes will be of interest for this umbrella review. Particular attention will be paid to the parameters used for measuring response to treatment in clinical settings with respect to established mechanism of actions of selenium. For example to measure the impact of selenium supplementation on patients suffering from trauma, indices such as incidence of nosocomial infection and hospital stay will be used^12^. While its effects on HIV-infected patients, outcomes measures such as CD4 count and viral suppression will be reported^15^

#### Type of studies

Only high quality systematic reviews with or without meta-analysis that included randomized controlled trials of selenium supplementation in disease conditions published from 2013 to present will be included in the review. While primary studies that involved the use of selenium supplementation alone will be excluded.

### Methods and analysis

The method of umbrella review in the manual for evidence synthesis published by JBI will be used^16^. The reporting method by the Preferred Reporting Items for Systematic Reviews and Meta-analyses (PRISMA) was used for writing this protocol and will be used for reporting the search results. In accordance with the guidelines, this umbrella review protocol was registered with PROSPERO the International Prospective Register of Systematic Reviews on 21^st^ March 2023 and last updated on 23^rd^ March 2023. The registration number is CRD42023406858. The JBI critical appraisal checklist will be used for the quality assessment.

#### Search strategy

Three reviewers will carry out the search using the strings containing the words derived from the title, abstract and keywords of this study (see Appendix 1). The following data bases will be searched: MEDLINE, PUBMED, EMBASE, COCHRANE database of systematic reviews, CINAHL, JBI Evidence synthesis, EPISTEMONIKOS, SCOPUS, Web of Science and TRIP PRO. Unpublished reviews will be searched using ProQuest for dissertations and Theses, Canadian Agency for drugs and Technologies in Health (CADTH) database of Grey matters and GOOGLE scholar. All systematic reviews including randomized clinical trials of selenium supplementation in sick patients published from 2013 to present. The reference lists for each of the identified systematic reviews will be searched as well for possible eligible studies. The number of results will be seen after importing the identified reviews into the Covidence. Duplicates will be removed before the starting the study selection phase.

#### Study selection

The study selection will be done by three independent reviewers by looking at the titles and abstracts of the results of the search strategy after the removal of duplicates. The eligibility criteria will be followed as the standard for appropriateness of choosing a particular study or not. The criteria include: systematic reviews that included controlled clinical trials of selenium alone in the management/treatment of disease condition in patients with clear measurable clinical outcomes, where there are updates on particular topic, the most recent updates will be chosen. At the end, the three reviewers will meet to compare the outcome of the selection process. Any disagreement will be resolved by consensus while uncertainties regarding the suitability of a study will be solved by including it for full-text review. Then, the selected references will be imported into Covidence and the three reviewers will carry out the full-text screening. The eligible articles will be moved on to the critical appraisal stage. Reasons for excluding any of the text will be recorded and reported in the PRISMA flowchart.

#### Assessment of methodological quality

The studies that were found appropriate after the full-text screening will be subjected to assessment of methodological quality by 3 independent assessors. The tool for this assessment will be the JBI critical appraisal checklist for systematic reviews and research syntheses^16^. A few examples of the questions in the appraisal tool are “Is the review question clearly and explicitly stated? Were the inclusion criteria appropriate for the review questions? For each of the JBI eleven quality criteria, the assessed will be assigned any three of the following responses: No/yes/Unclear/NA. No will be assigned 0 point, yes will be given 1 point. Each study will be classified as critically low quality if it has a score between 0-3, low quality for 4-6, moderate quality for 7-9 and high quality for 10-11. The overall quality of the studies will be used for the purposes of the umbrella review result interpretation and not for inclusion/exclusion of the selected systematic reviews. The results of the critical appraisal will be reported in a tabular form accompanied by narratives for easy interpretability.

#### Data collection

Extraction of data from the selected reviews will be done by 2 independent reviewers with the aid of the JBI data extraction tool ^16^ The data extraction will include the following study characteristics; information about the population, study objectives, context, publication date range and country of origin of the included studies, study location, study methods, outcome of interest relevant to the objectives of the review. In addition, other information will be extracted from the included reviews such as clinical outcomes, quantitative data on heterogeneity as well as results of meta-analysis (if done). Any disagreement will be resolved through discussion and a review by a third author if consensus was not reached. The data will be presented in a tabular format.

#### Data synthesis

A summary of the results from the critical appraisal of the included studies as well as the extracted data will be presented in tables. Then a narrative synthesis of the included reviews will be done to achieve the objectives of this umbrella review. If there are studies that were captured by more than one included systematic reviews, they will be clearly mentioned.

## Data Availability

All data produced in the present study are available upon reasonable request to the authors

## Ethics and dissemination

This study does not require ethical approval. The findings of the umbrella review will be presented in a conference as well as published in a peer-reviewed journal.

## Authors’ contributions

AUM is the guarantor. AUM, IU, AKA and BOU drafted the manuscript. AUM developed the search strategy. All authors contributed to the database selection, data extraction criteria. MOA and IU provided expertise on systematic reviews, the basis for the umbrella review. CSG provided insight on selenium supplementation. All authors read, provided feedback and approved the final manuscript.

## Funding statement

AUM is a beneficiary of Beca Guadalupe a scholarship by “Asociación Harambee” for a short stay at the University of Navarra, Spain towards acquisition of research skills needed to complete her PhD.

The funder does not have any role in the development of the protocol or conduct of this research.

AUM is a lecturer and doctoral fellow at the Department of Clinical Pharmacy and Pharmacy Management, University of Nigeria, Nsukka.

## Competing interests statement

The author declare no competing interests.

## Word count

The protocol contains 1470 words.

## Acknowledgement

University of Navarra, Pamplona Spain, for providing the access to some of the bibliographic databases.

María Marquínez Cabrejas from the Science library of the University of Navarra for her invaluable help starting from the fine-tuning of the research questions through the search strategy.

## Appendix I Search Strategy

### PubMed

Preliminary search conducted on March, 2 2023

| Search | Query | Records retrieved |
| --- | --- | --- |
| **#1** | "selenium"[MeSH Terms] OR "selenium"[All Fields] OR "seleniums"[All Fields] OR "seleniums"[All Fields] | 41,767 |
| **#2** | "systematic review"[Publication Type] OR "systematic reviews as topic"[MeSH Terms] OR "systematic reviews"[All Fields] | 256,226 |
| **#3** | "meta analysis"[Publication Type] OR "meta analysis as topic"[MeSH Terms] OR "meta analysis"[All Fields] | 268,430 |
| **#2 OR #3 AND #3** | ("meta analysis"[Publication Type] OR "meta analysis as topic"[MeSH Terms] OR "meta analysis"[All Fields] OR ("systematic review"[Publication Type] OR "systematic reviews as topic"[MeSH Terms] OR "systematic reviews"[All Fields])) AND ("selenium"[MeSH Terms] OR "selenium"[All Fields] OR "seleniums"[All Fields] OR "seleniums"[All Fields]) | 464 |
| **Date filter (2013 to present)** |  | 349 |

## Notes

### Competing Interest Statement

The authors have declared no competing interest.

### Funding Statement

This study did not receive any funding

